# Epidemiological patterns and factors associated with mortality among diphtheria cases in Kano State, Nigeria (2022-2025): A retrospective cohort study

**DOI:** 10.64898/2026.09.22.26363747

**Authors:** Paul Waliaula Wekunda, Oladipo Ogunbode, Oyeladun Okunromade

## Abstract

**Introduction:** Nigeria continues to report one of the highest burdens of diphtheria globally, with Kano State accounting for a disproportionate share of cases and deaths. Understanding epidemiological patterns and mortality determinants is essential for guiding targeted interventions.

**Objectives:** To describe the distribution of diphtheria cases in Kano State by socio-demographic and clinical characteristics, Local Government Area, and month of reporting, and identify independent predictors of mortality.

**Methods:** We conducted a retrospective cohort study of 21,369 patients with clinically compatible and laboratory-confirmed diphtheria using secondary data from medical records (2022–2025). Descriptive statistics summarized patient characteristics and disease distribution. Associations with mortality were assessed using chi-square or Fisher’s exact tests. Multivariable logistic regression identified predictors of mortality, reported as adjusted odds ratios (aORs) and 95% confidence intervals (CIs).

**Results:** Of 21,369 patients, 13,557 (63%) were female, with a median age of 11 years (IQR: 7–13.6). Children aged 5–9 years accounted for 29% of cases. Prior contact with a case was reported in 2,633 (12%) patients, while 168 (1%) were laboratory-confirmed. Overall, 13,468 (63%) patients were unvaccinated and 5,381 (25%) fully vaccinated; Diphtheria antitoxin DAT was administered to 2,035 (10%). Cases occurred in all local government areas (LGAs), with four accounting for 60% of cases. Transmission occurred year-round, peaking in March 2024 and August 2023 and 2025. Overall, 1,131 deaths occurred (CFR: 5.3%). Compared with patients aged ≥20 years, younger age groups had higher odds of mortality: 0–4 years (aOR = 26.74, 95% CI: 17.64–42.72), 5–9 years (aOR = 19.77, 95% CI: 13.17–31.33), 10–14 years (aOR = 8.21, 95% CI: 5.41–13.13), and 15–19 years (aOR = 2.46, 95% CI: 1.45–4.26). Prior contact with a case was associated with lower odds of mortality (aOR = 0.42, 95% CI: 0.30–0.55), whereas laboratory confirmation (aOR = 2.33, 95% CI: 1.48–3.56) and DAT receipt (aOR = 2.55, 95% CI: 2.19–2.96) were associated with higher odds. Prior vaccination was associated with lower odds of mortality among fully vaccinated (aOR = 0.52, 95% CI: 0.44–0.61) and partially vaccinated patients (aOR = 0.61, 95% CI: 0.47–0.80).

**Conclusions and recommendations:** During the study period, diphtheria was more common among younger children, females, and the unvaccinated. Although concentrated in a few LGAs, transmission was widespread and varied seasonally. Mortality was associated with younger age, laboratory-confirmed infection, and use of diphtheria antitoxin, whereas vaccination and prior contact were protective. Reducing the burden requires sustained efforts to strengthen routine immunization, implement targeted vaccination campaigns, ensure early detection and prompt treatment, and improve contact tracing and case management.

## Introduction

Diphtheria is a life-threatening vaccine-preventable infectious disease caused by *Corynebacterium diphtheriae*, a gram-positive, rod-shaped bacterium (1). Transmission occurs primarily through respiratory droplets, although contact with contaminated surfaces may also contribute to the spread. After infection, the organism produces a potent exotoxin that binds to host cells in the respiratory tract, heart, skin, kidneys, and nervous system, causing cellular damage and necrosis (2). The respiratory form is the most common clinical presentation. It is characterized by a greyish pseudomembrane over the throat and tonsils, which can cause airway obstruction and difficulty swallowing (3). Cutaneous diphtheria may also occur, typically presenting as chronic skin lesions or ulcers (4). While the disease can affect individuals of all age groups, unvaccinated children are particularly at risk (5–7)

Despite substantial progress in global diphtheria prevention and control, significant systemic challenges persist in low- and middle-income countries, where the disease remains an important public health threat (5,8). Nigeria is among the countries with the highest reported diphtheria case counts globally. The resurgence of the disease in this setting has been driven by multiple interconnected factors, including gaps in vaccination coverage, weak health systems, low socioeconomic status, limited access to healthcare services, insecurity, and disruptions associated with the COVID-19 pandemic (9–13). Between epidemiological week (Epi-week) 19 of 2022 and Epi-week 18 of 2025, Nigeria reported 43,743 suspected diphtheria cases across all 36 states and the Federal Capital Territory (FCT) (14). Of these, 26,499 were clinically compatible and laboratory-confirmed, with 1,376 associated deaths, yielding a case fatality ratio (CFR) of 5.2%. Kano State bears the highest burden, accounting for 24,415 suspected and 18,384 confirmed cases, along with a disproportionately high number of deaths.

However, there was limited evidence on the geographical and temporal distribution of cases, as well as on factors associated with mortality in the high-burden setting. Understanding these patterns is essential for informing targeted public health interventions. Therefore, this study aimed to describe the distribution of diphtheria cases in Kano State by socio-demographic and clinical characteristics, local government area (LGA), and month of reporting, and to identify independent predictors of mortality between 2022 and 2025.

## Methods

### Study setting

This study was conducted in Kano State, located in northern Nigeria. The state was purposively selected due to its consistently high burden of diphtheria cases. Kano State comprised 44 LGAs and had an estimated population of approximately 16.3 million in 2024, making it the most populous state in Nigeria (15).

### Study design

A retrospective cohort study was conducted over four years, from September 2022 to November 2025, among patients diagnosed with diphtheria.

### Study participants and sample size

The study included all patients diagnosed with diphtheria and classified as either clinically compatible or laboratory confirmed. Patients from other states and those with unclear case classification were excluded. The analysis utilized secondary data on 21,369 patients, with an observed mortality rate of approximately 5%. Given the fixed dataset and outcome (mortality) frequency, a priori sample size calculation was not performed.

### Data collection

The secondary data were obtained through a review of medical records for diphtheria patients reported between 2022 and 2025. It was case-based data, containing various variables for each patient.

### Study variables

The dependent variable for this study was the outcome at discharge (dead or alive). Predictor variables included socio-demographic characteristics, sex/gender (male or female), age (categorized into age groups), and history of prior contact with a diphtheria case (yes, no), as well as clinical factors such as case classification (clinically compatible or laboratory confirmed), vaccination status (unvaccinated, fully vaccinated, partially vaccinated, or unknown), receipt of diphtheria antitoxin (yes or no), and hospitalization status (yes or no). Additional variables included presenting clinical features, geographic location (LGA), and time of reporting (month and year).

## Data analysis

Data was analyzed using R. Descriptive statistics, including frequencies, proportions, and medians with interquartile ranges (IQRs), summarized socio-demographic and clinical characteristics, as well as the geographic and temporal distribution of cases. Bivariate analyses using the Chi-square test or Fisher’s exact test, as appropriate, assessed associations between categorical variables and mortality. We performed multivariable logistic regression with epidemiologically relevant variables to identify independent predictors of mortality. Results were presented as adjusted odds ratios (aORs) with 95% confidence intervals (CIs). We evaluated logistic regression model fit using the Hosmer–Lemeshow goodness-of-fit test, with a p-value >0.05 indicating adequate fit (16).

## Ethical consideration

We obtained ethics approval from the Kano State Ministry of Health Research Ethics Committee (Ethics clearance reference SHREC/2026/7812). Surveillance data were accessed on June 6, 2026. Data privacy, confidentiality, and anonymity were strictly maintained throughout the study to protect patients’ sensitive information.

## Results

Between September 2022 and November 2025, a total of 21,369 clinically compatible and laboratory-confirmed diphtheria cases were reported in Kano State. The median age was 11 years (Interquartile Range [IQR]: 7–13.6 years). Overall, 1,131 patients died, resulting in a cumulative mortality incidence of 5.3%.

### Demographic and Clinical Characteristics of Patients Diagnosed with Diphtheria in Kano State, Nigeria

Table 1 presents the baseline demographic and clinical characteristics of patients with diphtheria in Kano State. Of the 21,369 cases, 13,557 (63%) were female. Children aged 5–9 years accounted for 6,108 (29%) cases, while those aged 10–14 years accounted for 5,708 (27%). Regarding exposure history, 2,633 (12%) patients reported known previous contact with a diphtheria case. Laboratory confirmation was available for 168 (1%) cases. In terms of vaccination status, 13,468 (63%) patients had never been vaccinated against diphtheria, whereas 5,381 (25%) were fully vaccinated. Overall, 2,035 (10%) patients received diphtheria antitoxin (DAT), including 56 (33%) of laboratory-confirmed cases and 1,979 (9.3%) of clinically compatible cases. Adverse events following DAT administration were reported in two cases. A total of 8,800 (41%) patients were hospitalized, comprising 155 (92%) laboratory-confirmed cases and 8,645 (41%) clinically compatible cases.

**Table 1:**
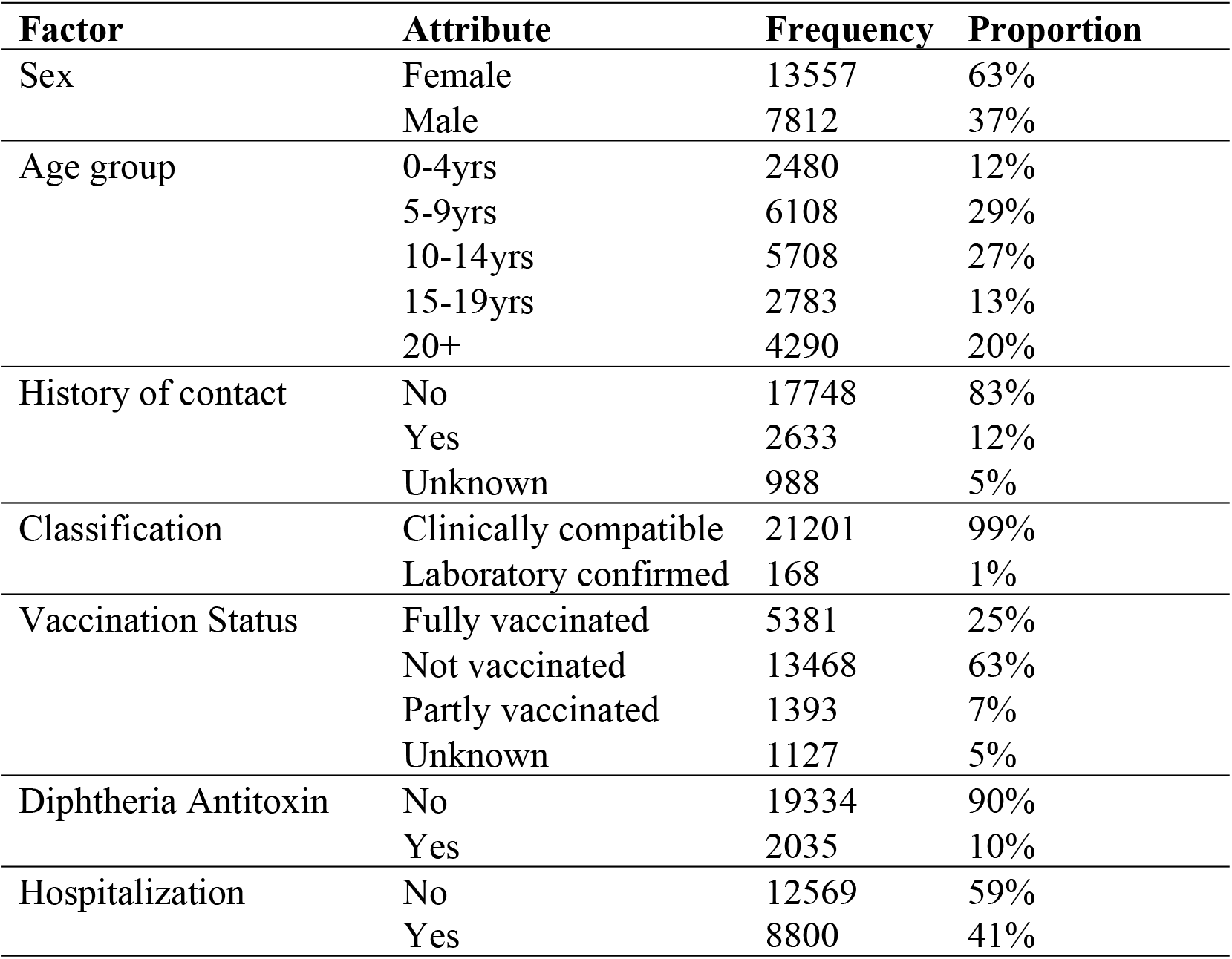
Demographic and Clinical Characteristics of Patients Diagnosed with Diphtheria in Kano State, 2022-2025.

Table 2 highlights the clinical features of diphtheria patients in Kano State, based on symptoms reported by patients and/or findings observed by clinicians. All patients presented with a diphtheria-related pseudomembrane. Other common clinical features included pharyngitis (98%), tonsillitis (97%), and fever (94%). In contrast, cough occurred in 854 (4%) patients, while none presented with diphtheria-related skin lesions.

**Table 2:** Clinical features of patients diagnosed with diphtheria in Kano State, 2022-2025.

| Symptom | Attribute | Frequency | Proportion |
| --- | --- | --- | --- |
| Pseudomembrane | Yes | 21369 | 100% |
|  | No | 0 | 0% |
| Pharyngitis | Yes | 20870 | 98% |
|  | No | 499 | 2% |
| Tonsillitis | Yes | 20803 | 97% |
|  | No | 566 | 3% |
| Fever | Yes | 19993 | 94% |
|  | No | 1376 | 6% |
| Neck Swelling | Yes | 2869 | 13% |
|  | No | 18500 | 87% |
| Laryngitis | Yes | 2695 | 13% |
|  | No | 18404 | 86% |
| Nasopharyngitis | Yes | 907 | 4% |
|  | No | 20462 | 96% |
| Cough | Yes | 854 | 4% |
|  | No | 20515 | 96% |
| Skin lesion | Yes | 16 | 0% |
|  | No | 21353 | 100% |
| Outcome | Alive | 20238 | 95% |
|  | Dead | 1131 | 5% |

### Distribution of diphtheria cases in Kano State by Local Government Areas and months (2022 and 2025)

The distribution of diphtheria cases by LGA in Kano State is presented in Figure 1. All LGAs reported cases of diphtheria during the study period. Of the 21,369 total reported cases, Ungogo LGA accounted for 5,791 cases (27%). Overall, four LGAs, Ungogo, Dala, Fagge, and Kumbotso, collectively accounted for 12,761 (60%) of all reported cases in Kano State.

**Figure 1:**
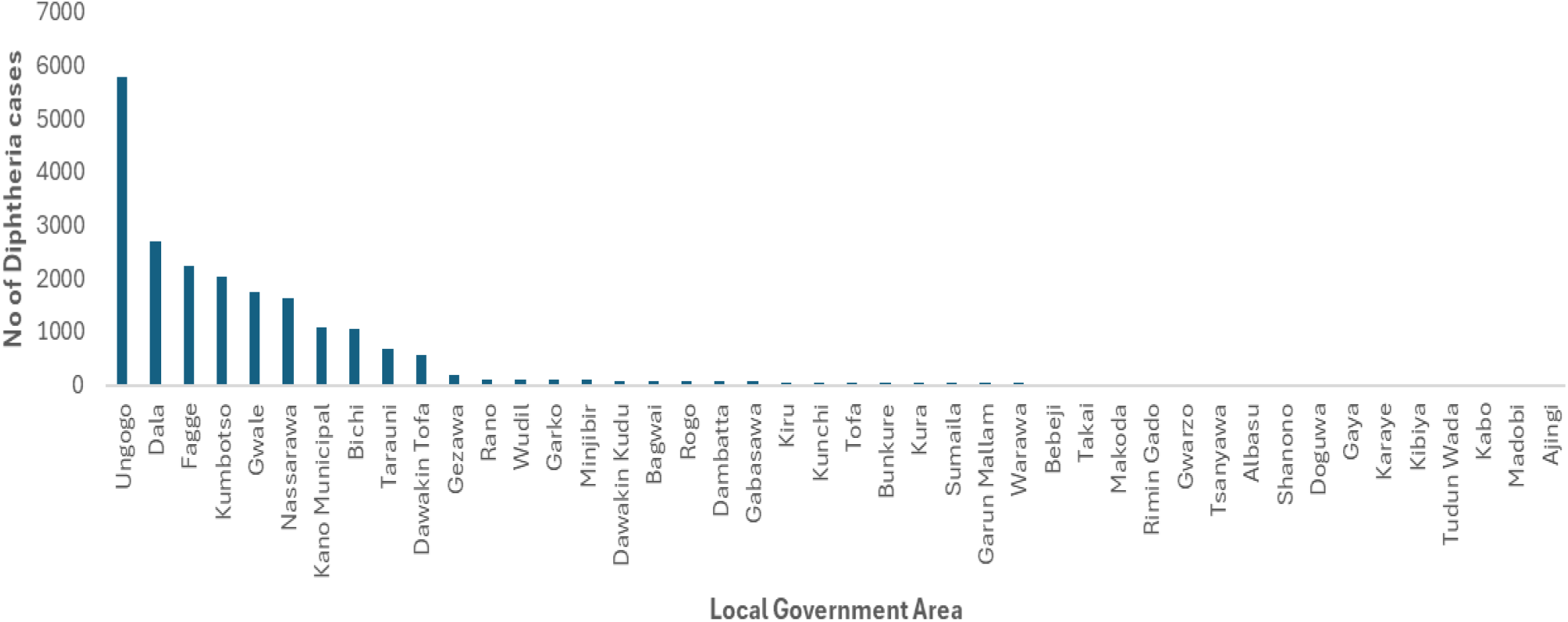
Distribution of diphtheria cases in Kano State by Local Government Areas, 2022-2025

Figure 2 illustrates the temporal trend of diphtheria cases in Kano State from 2022 to 2025. Case reporting began in September 2022, and cases were reported each month consistently throughout the study period. The highest number of cases was recorded in 2023, with 10,518 cases (49% of the total). In 2024, cases peaked in March, whereas in both 2023 and 2025, peak transmission occurred in August.

**Figure 2:**
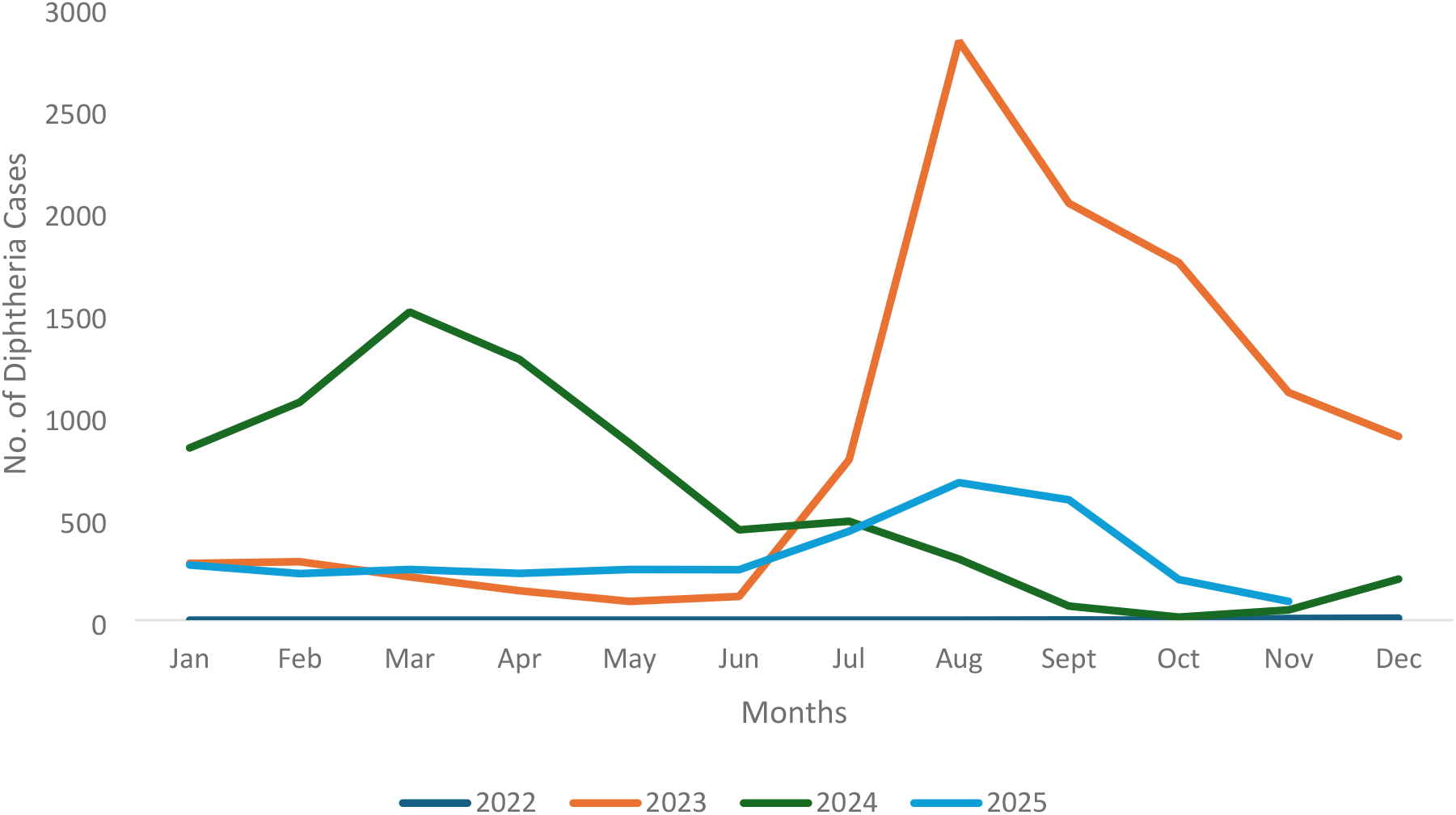
Temporal trend of diphtheria cases in Kano State from 2022 to 2025

### Association between demographic and clinical factors and mortality among patients with diphtheria

A chi-square test of independence was conducted to examine bivariate associations between selected demographic and clinical characteristics and mortality, as presented in Table 3. There was a statistically significant association between patient sex and mortality, χ^2^ (1, N = 21,369) = 16.51, p < .001. Age group was also significantly associated with mortality, χ^2^ (4, N = 21,369) = 689.34, p < .001. Similarly, a significant association was observed between prior contact with a diphtheria case and mortality, χ^2^ (2, N = 21,369) = 74.61, p < .001. Case classification showed a significant relationship with outcome, χ^2^ (2, N = 21,369) = 41.45, p < .001, with laboratory-confirmed cases demonstrating a higher likelihood of mortality compared to clinically compatible cases. Vaccination status was also significantly associated with mortality, χ^2^ (3, N = 21,369) = 31.76, p < .001. Finally, receipt of DAT and hospitalization were both associated with higher mortality.

**Table 3:** Association between demographic and clinical factors and mortality among patients with diphtheria, 2022-2025.

| Factor | Attribute | Frequency | Number Died (%) | Chi-square (p-value) |
| --- | --- | --- | --- | --- |
| Sex | Female | 13557 | 653 (5%) | 16.51 (<0.001) |
|  | Male | 7812 | 478 (6%) |  |
| Age group | 0-4yrs | 2480 | 296 (12%) | 689.34 (<0.001) |
|  | 5-9yrs | 6108 | 549 (9%) |  |
|  | 10-14yrs | 5708 | 229 (4%) |  |
|  | 15-19yrs | 2783 | 35 (1%) |  |
|  | 20+ | 4290 | 22 (1%) |  |
| History of contact | No | 17748 | 1045 (6%) | 74.61 (<0.001) |
|  | Yes | 2633 | 58 (2%) |  |
|  | Unknown | 988 | 28 (3%) |  |
| Classification | Clinically compatible | 21201 | 1103 (5%) | 41.45 (<0.001) |
|  | Laboratory confirmed | 168 | 28 (17%) |  |
| Vaccination Status | Not vaccinated | 13468 | 801 (6%) | 31.76 (<0.001) |
|  | Fully vaccinated | 5381 | 219 (4%) |  |
|  | Partially vaccinated | 1393 | 64 (5%) |  |
|  | Unknown | 1127 | 47 (4%) |  |
| Diphtheria Antitoxin | No | 19334 | 848 (4%) | 331.04 (<0.001) |
|  | Yes | 2035 | 283 (14%) |  |
| Hospitalization | No | 12569 | 121 (1%) | 1139.5 (<0.001) |
|  | Yes | 8800 | 1010 (11%) |  |

The Hosmer–Lemeshow goodness-of-fit test was performed to assess logistic regression model fit and indicated an adequate fit (χ^2^ = 8.12, p = 0.42). With over 1,131 deaths observed, the study had sufficient statistical power to support multivariable analysis and detect clinically meaningful associations.

Table 4 presents the results of both univariable and multivariable logistic regression analyses conducted to identify independent predictors of mortality. Compared to patients aged ≥20 years, those in younger age groups had higher odds of mortality: 0–4 years (adjusted odds ratio [aOR] = 26.74, 95% confidence interval [CI]: 17.64–42.72), 5–9 years (aOR = 19.77, 95% CI: 13.17–31.33), 10–14 years (aOR = 8.21, 95% CI: 5.41–13.13), and 15–19 years (aOR = 2.46, 95% CI: 1.45–4.26). Patients with a history of prior contact with a diphtheria case had significantly lower odds of mortality compared to those without such contact (aOR = 0.42, 95% CI: 0.30–0.55). In contrast, laboratory-confirmed cases had higher odds of mortality compared to clinically compatible cases (aOR = 2.33, 95% CI: 1.48–3.56). Vaccination status was also significantly associated with mortality. Compared with unvaccinated patients, prior vaccination was associated with lower odds of mortality among fully vaccinated (aOR = 0.52, 95% CI: 0.44–0.61) and partially vaccinated patients (aOR = 0.61, 95% CI: 0.47–0.80). Finally, patients who received DAT had higher odds of mortality compared to those who did not (aOR = 2.55, 95% CI: 2.19–2.96).

**Table 4:** Predictors of mortality among diphtheria patients, 2022-2025.

| Factor | Attribute | Frequency | Number Died (%) | Crude Odds ratio (95% CI) | Adjusted Odds ratio (95% CI) |
| --- | --- | --- | --- | --- | --- |
| Sex | Female | 13557 | 653 (5%) | Ref |  |
|  | Male | 7812 | 478 (6%) | 1.29 (1.14,1.46) | 0.91 (0.8,1.03) |
| Age group | 20+ | 4290 | 22 (1%) | Ref |  |
|  | 0-4yrs | 2480 | 296 (12%) | 26.29 (17.42,41.86) | 26.74 (17.64,42.72) |
|  | 5-9yrs | 6108 | 549 (9%) | 19.16 (12.81,30.29) | 19.77 (13.17,31.33) |
|  | 10-14yrs | 5708 | 229 (4%) | 8.11 (5.35,12.94) | 8.21 (5.41,13.13) |
|  | 15-19yrs | 2783 | 35 (1%) | 2.47 (1.46, 4.28) | 2.46 (1.45,4.26) |
| History of contact | No | 17748 | 1045 (6%) | Ref |  |
|  | Yes | 2633 | 58 (2%) | 0.36 (0.27,0.47) | 0.42 (0.32,0.55) |
|  | Unknown | 988 | 28 (3%) | 0.47 (0.31,0.67) | 0.6 (0.39,1) |
| Classification | Clinically compatible | 21201 | 1103 (5%) | Ref |  |
|  | Laboratory confirmed | 168 | 28 (17%) | 3.64 (2.37,5.4) | 2.33 (1.48,3.56) |
| Vaccination Status | Not vaccinated | 13468 | 801 (6%) | Ref |  |
|  | Fully vaccinated | 5381 | 219 (4%) | 0.67 (0.57,0.78) | 0.52 (0.44,0.61) |
|  | Partially vaccinated | 1393 | 64 (5%) | 0.76 (0.58,0.98) | 0.61 (0.47,0.8) |
|  | Unknown | 1127 | 47 (4%) | 0.68 (0.5,0.92) | 0.91 (0.66,1.23) |
| Diphtheria Antitoxin | No | 19334 | 848 (4%) | Ref |  |
|  | Yes | 2035 | 283 (14%) | 3.52 (3.04,4.07) | 2.55 (2.19,2.96) |

## Discussion

This study characterized the epidemiology of diphtheria and identified factors associated with mortality in Kano State between 2022 and 2025. The findings indicate a substantial disease burden, marked by high mortality and widespread distribution across all Local Government Areas (LGAs). Four LGAs, Ungogo, Dala, Fagge, and Kumbotso, accounted for 60% of reported cases, highlighting localized transmission hotspots that may benefit from targeted public health interventions (13). The temporal pattern suggests sustained year-round transmission with variable seasonal peaks. While incidence peaked in March in 2024, peaks occurred in August in both 2023 and 2025, indicating inconsistent seasonality. This variability likely reflects the influence of multiple contextual factors on transmission dynamics, including substandard living conditions, asymptomatic carriers, overcrowding, conflict and insurgency, and low immunization coverage (7,9,11,17–19). Overall, these findings underscore the need for continuous, rather than seasonally focused, prevention and control strategies, alongside interventions that address broader social and structural determinants of health.

Clinically, the universal presence of pseudomembrane and the high prevalence of pharyngitis, tonsillitis, and fever are consistent with the classical presentation of diphtheria (20–22). The low frequency of cough and absence of skin lesions suggest that upper respiratory diphtheria (6) was the predominant clinical form in Kano State.

Although a higher proportion of deaths occurred among males, females accounted for approximately 63% of cases, indicating a greater overall disease burden among females. Similar sex distributions have been reported in previous studies (23–25). In contrast, a study conducted in Indonesia (26) found a higher prevalence among males, possibly reflecting contextual differences in exposure and inherent risk factors. This study revealed that 57% of cases occurred among individuals younger than 14 years, consistent with findings from studies within and outside Nigeria (20,25,27). Mortality was disproportionately higher among younger age groups, aligning with existing literature (21,28). This increased vulnerability may be attributed to incomplete vaccination, immature or waning immune responses, and delays in accessing appropriate care (19,29). Strengthening routine immunization and ensuring timely booster doses in childhood may therefore reduce preventable deaths.

Interestingly, patients with a known history of contact with a diphtheria case had a 58% lower risk of mortality compared to those without known exposure. This apparent protective effect likely reflects earlier identification through contact tracing and more timely care-seeking among known contacts, rather than a true biological advantage. Previous studies suggest that contact tracing facilitates early symptom recognition, enabling timely diagnosis and prompt initiation of treatment (9,30,31). In addition, heightened awareness among contacts, along with community and peer support, may contribute to improved care-seeking behavior and better clinical outcomes.

The high proportion of unvaccinated individuals among reported cases, together with the observed protective effect of vaccination against mortality, underscores the important role of immunization in preventing diphtheria and reducing disease severity. Similar patterns of low vaccination coverage among diphtheria cases have been reported in other studies in Nigeria (9,18,21). Because diphtheria-containing vaccines are administered through pentavalent and tetanus–diphtheria formulations, gaps in vaccination coverage may increase susceptibility not only to diphtheria but also to other vaccine-preventable diseases. Strengthening immunization strategies, particularly in high-burden LGAs, is therefore important for improving population immunity and reducing preventable morbidity and mortality. This requires sustained investment in immunization programs, strengthened disease surveillance, and greater collaboration among relevant stakeholders (12). However, the occurrence of cases among fully vaccinated individuals may reflect waning immunity, highlighting the potential importance of booster doses, particularly among populations at increased risk (32–34). In addition, previous studies have suggested that vaccination alone may interrupt transmission in only 28% of outbreak settings, emphasizing the complementary importance of case isolation and appropriate antibiotic treatment in controlling transmission (13).

The observed association between higher mortality and both diphtheria antitoxin (DAT) administration and laboratory confirmation is likely attributable to confounding by indication. A substantially higher proportion of laboratory-confirmed cases were hospitalized than clinically compatible cases (92% vs. 41%) and received DAT more frequently (33% vs. 9.3%). This pattern suggests that patients with more severe disease were more likely to undergo confirmatory testing and receive DAT, while also being at inherently higher risk of mortality. Therefore, these associations should not be interpreted as causal but rather as potential proxies for disease severity at presentation. Similar findings were reported by Denue *et al*. (28), whereas Alege *et al*. (21) reported contrasting results, possibly reflecting differences in the timing of DAT administration. Although DAT is essential for neutralizing circulating diphtheria toxin, it cannot neutralize toxins that are already bound to tissues (2,35,36). This underscores the importance of early DAT administration alongside appropriate antibiotic therapy. Given the critical role of DAT in diphtheria management, further studies are warranted to clarify its relationship with mortality, particularly by examining the timing of treatment initiation, disease severity at presentation, and health-system factors that may influence patient outcomes.

## Conclusion

This study demonstrates that despite being a vaccine-preventable disease, diphtheria remains endemic in Kano State, disproportionately affecting younger individuals, females, and those who are unvaccinated. While cases were concentrated in a few high-burden LGAs, transmission was widespread across the state, occurring year-round with variable seasonal peaks. Predictors of mortality included younger age, laboratory-confirmed infection, and receipt of DAT, whereas prior vaccination (full or partial) and a history of contact with a confirmed case appeared to confer protective effects.

Reducing the burden of diphtheria in Kano State will require sustained and coordinated public health interventions. These should include strengthening routine and booster vaccination programs, implementing targeted vaccination campaigns in high-risk areas, ensuring early case detection and prompt case management, particularly among children, and enhancing effective contact tracing. In addition, robust community engagement is essential to improve public awareness, increase vaccine uptake, and promote timely healthcare-seeking behavior.

### Strengths and Limitations of the Study

This study benefited from a large sample size and multi-year data covering all LGAs in Kano State, enhancing the internal generalizability of the findings. Additionally, multivariable analysis identified independent predictors of mortality. However, several limitations should be considered. Key clinical variables, such as the timing of treatment initiation, disease severity at presentation, and the presence of comorbidities, were unavailable, limiting the ability to fully account for confounding. Notably, the associations observed with diphtheria antitoxin administration and laboratory confirmation are likely influenced by unmeasured indicators of disease severity.

## Data Availability

All data underlying the findings of this study have been deposited in the Figshare and can be accessed at 10.6084/m9.figshare.33944044 under the accession number CC.BY. 40

## References

1. Guilfoile P. Diphtheria [Internet]. New York; 2009. 1–119 p. Available from: https://books.google.com.ng/books?hl=en&lr=&id=tzVqM-JUnW8C&oi=fnd&pg=PP1&dq=diphtheria+causes&ots=GByxe6mzML&sig=N88t-zay1I316XLya55hmjG0uG4&redir_esc=y#v=onepage&q=diphtheriacauses&f=false

2. Grandière Pérez L, Brisse S. Diphtheria antitoxin treatment: from pioneer to neglected. Mem Inst Oswaldo Cruz. 2025;119:e240214. doi:10.1590/0074-02760240214 PubMed PMID: 39841756.

3. Forde BM, Henderson A, Playford EG, Looke D, Henderson BC, Watson C, et al. Fatal Respiratory Diphtheria Caused by ß-Lactam – Resistant Corynebacterium diphtheriae. Clinical Infectious Disease. 2021;73(11):e4531–8. doi:10.1093/cid/ciaa1147

4. Chêne L, Morand JJ, Badell E, Toubiana J, Janvier F, Marthinet H, et al. Cutaneous diphtheria from 2018 to 2022: an observational, retrospective study of epidemiological, microbiological, clinical, and therapeutic characteristics in metropolitan France. Emerg Microbes Infect. 2024;13(1). doi:10.1080/22221751.2024.2408324 PubMed PMID: 39324172.

5. Wu Z, Lin L, Zhang J, Zhong J, Lai D. Global burden of diphtheria, 1990–2021: a 204-country analysis of socioeconomic inequality based on SDI and DTP3 vaccination differences before and after the COVID-19 pandemic (GBD 2021). Front Public Health. 2025;13:1597076. doi:10.3389/fpubh.2025.1597076 PubMed PMID: 40620556.

6. Kęder K, Kukulska MB, Latocha J, Sobieska A, Grzelak D. Diphtheria-Epidemiology situation, pathogenesis, diagnosis, treatment methods and prevention. Przeglad epidemiologiczny. 2025. p. 215–26. doi:10.32394/pe/208640 PubMed PMID: 41042958.

7. Johnson OS, Edogbanya HO, Wakili A, John AT. Diphtheria Disease Transmission Dynamics in Low Vaccine Coverage Setting. International Journal of Mathematical Sciences and Optimization: Theory and Applications. 2024;10(2):79–106. doi:10.5281/zenodo.10966277

8. Clarke KEN, Macneil A, Hadler S, Scott C, Tiwari TSP, Cherian T. Global Epidemiology of Diphtheria, 2000–2017. Emerg Infect Dis. 2019;25(10):1834–42. doi:10.3201/eid2510.190271 PubMed PMID: 31538559.

9. Oduoye MO, Dheyaa M, Marsool M, Haider MU, Karim KA. Unmasking diphtheria in Nigeria: A multifaceted approach to tackle outbreaks and improve immunization rates among the Nigerian population — An updated correspondence. Health Sci Rep. 2024;7:e1804. doi:10.1002/hsr2.1804

10. Osarenren J, Omoruyi P, Olalekan O, Okesanya J, Omosigho PO, Olalekan JO. Global strategies for addressing diphtheria resurgence, epidemiology, clinical impact, and prevention. Discover Public Health. 2024;21(219). doi:10.1186/s12982-024-00352-1

11. Abbas MA, Yusuf AL, Murtala HA, Abdullahi AA, Murtala AM, Torrelles JB, et al. Post-COVID-19 resurgence of diphtheria in Kano, Nigeria: Analysis of 18,320 cases. EBioMedicine. 2025;118:105877. doi:10.1016/j.ebiom.2025.105877

12. Osarenren J, Omosigho OP, Okesanya OJ. Global strategies for addressing diphtheria resurgence, epidemiology, clinical impact, and prevention. Discover Public Health. 2024;21(219). doi:10.1186/S12982-024-00352-1

13. Truelove SA, Keegan LT, Moss WJ, Chaisson LH, Macher E, Azman AS, et al. Clinical and Epidemiological Aspects of Diphtheria: A Systematic Review and Pooled Analysis. Clinical Infectious Diseases. 2020 Jun 24;71(1):89–97. doi:10.1093/CID/CIZ808 PubMed PMID: 31425581.

14. Nigeria Centre for Disease Control and Prevention. Weekly diphtheria situation report: As of 3rd May 2025 (Epi-week 18, 2025) [Internet]. Abuja; 2025. Available from: https://ncdc.gov.ng/diseases/sitreps/?cat=18&name=AnUpdateofDiphtheriaOutbreakinNigeria

15. National Population Commision. Nigeria 2024 Demographic and Health Survey: Summary Report. 2024.

16. Surjanovic N, Loughin TM. Improving the Hosmer-Lemeshow goodness-of-fit test in large models with replicated Bernoulli trials. Journal of Applied Statistics. Taylor and Francis Ltd.; 2024. p. 1399–411. doi:10.1080/02664763.2023.2272223

17. Oduoye MO, Musa ZM, Tunde AM, Nazir A, Cakwira H, Abdulkareem L, et al. The recent outbreak of diphtheria in Nigeria is a public health concern for all. International Journal of Surgery: Global Health. 2023;6(5). doi:10.1097/gh9.0000000000000274

18. Abdulrasheed N, Lawal L, Mogaji AB, Abdulkareem AO, Shuaib AK, Adeoti SG, et al. Recurrent diphtheria outbreaks in Nigeria: A review of the underlying factors and remedies. Immun Inflamm Dis. 2023;11:e1096. doi:10.1002/iid3.1096 PubMed PMID: 38018582.

19. Olulaja ON, Anjorin ET, Ekerin O, Afolabi OT, Inuojo JM. A looming epidemic: combating the recurrent outbreaks of diphtheria in Nigeria. Pan Afr Med J. 2023;45(186). doi:10.11604/pamj.2023.45.186.41328 PubMed PMID: 38020360.

20. Zulfan GP, Sihombim AJ, Desrinawati MA, Widiantari DA, Berti PEM, Murtiani F. Clinical Manifestation of Childhood Diphtheria. Jurnal Ilmiah Kedokteran Wijaya Kusuma. 2023;12(1):1–6.

21. Alege A, Ibrahim OR, Ibraheem RM, Aladesua O, Lugga AS, Yahaya YY, et al. Clinical presentation and predictors of hospital mortality of diphtheria in Nigeria, July 2023 to April 2024: a single-center study. BMC Infect Dis. 2025;25(8). doi:10.1186/s12879-024-10401-4 PubMed PMID: 39748294.

22. Erim A, Lawal HO, Onyeaghala CA, Chukwu-Mba CC, Ajibade OO, Okafor UG, et al. Seasonal variations in public perceptions of diphtheria in Northern Nigeria. BMC Public Health. 2025;25(2146). doi:10.1186/s12889-025-23427-3 PubMed PMID: 40495171.

23. Shedaiwah S, Alsharabi H, Anam L, Al Amad MA. Risk factors of diphtheria outbreak in damt district of Al Dhalea Governorate, 2023 -Yemen: a case–control study. BMC Infect Dis. 2024;24(1034). doi:10.1186/s12879-024-09932-7 PubMed PMID: 39333947.

24. Moulhee NMS, Alwesabi SAM, Taha MS, Majam ASM. Epidemiological Analysis of Diphtheria Cases in Hodeida, Yemen: A Review of Admission Records from 2018 to 2022. Journal of Medical and Pharmaceutical Sciences. 2025;9(4):29–37. doi:10.26389/ajsrp.b180825

25. Ikejezie J, Langley T, Lewis S, Bisanzio D, Phalkey R. The epidemiology of diphtheria in Haiti, December 2014–June 2021: A spatial modeling analysis. PLoS One. 2022;17(8):e0273398. doi:10.1371/journal.pone.0273398 PubMed PMID: 35994502.

26. Husada D, Hartini Y, Nuringhati KW, Tindage SG, Mustikasari RI, Kartina L, et al. Eleven-Year Report of High Number of Diphtheria Cases in Children in East Java Province, Indonesia. Trop Med Infect Dis. 2024;9(9). doi:10.3390/tropicalmed9090204

27. Ibrahim OR, Lawal IM, Mohammed B, Abdullahi SB, Bello SO, Issa A, et al. Diphtheria outbreak during Covid-19 pandemic in Katsina, North-Western Nigeria: Epidemiological characteristics and predictors of death. Nigerian Journal of Basic and Clinical Sciences. 2022;19:59–65. doi:10.4103/njbcs.njbcs_35_21

28. Denue BA, Ngoshe RM, Abdul H, Akawu CB, Gana ML, Hussaini AY, et al. Clinical presentation and outcome of diphtheria in health facility in North-East Nigeria. Egypt J Intern Med. 2024;36(105). doi:10.1186/s43162-024-00372-y

29. Muscat M, Gebrie B, Efstratiou A, Datta SS, Daniels D. Diphtheria in the WHO European Region, 2010 to 2019. Eurosurveillance. 2022;27(8):2100058. doi:10.2807/1560-7917.ES.2022.27.8.2100058 PubMed PMID: 35209973.

30. World Health Organization. WHO Guidelines on Contact Tracing. Geneva; 2024.

31. Djaafara BA, Adrian V, Eriawati E, Elyazar IRF, Hamers RL, Baird JK, et al. Modeling the transmission dynamics and control strategies during the 20ti17 diphtheria outbreak in Jakarta, Indonesia. Infect Dis Model. 2026 Mar 1;11(1):1–15. doi:10.1016/j.idm.2025.08.004

32. Truelove SA, Keegan LT, Moss WJ, Chaisson LH, Macher E, Azman AS, et al. Clinical and epidemiological aspects of diphtheria: A systematic review and pooled analysis. Clinical Infectious Diseases. 2020;71(1):89–97. doi:10.1093/cid/ciz808 PubMed PMID: 31425581.

33. Vusirikala A, Tonge S, Bell A, Linley E, Borrow R, O’Boyle S, et al. Reassurance of population immunity to diphtheria in England: Results from a 2021 national serosurvey. Vaccine. 2023;41(46):6878–83. doi:10.1016/j.vaccine.2023.10.003 PubMed PMID: 37821313.

34. Murhekar M V, Kamaraj P, Kumar MS, Khan SA, Allam RR, Barde P V. Immunity against diphtheria among children aged 5–17 years in India, 2017–18: a cross-sectional, population-based serosurvey. Lancet Infectious Diseases. 2021;21(6):868–75. doi:10.1016/S1473-3099(20)30595-8

35. Anish Lamichhane, Sajithkumar Radhakrishnan. Diphtheria. In: Manson’s Tropical Diseases, Fourth Edition [Internet]. StatPearls Publishing; 2024 [cited 2026 Apr 17]. p. 461–5. Available from: https://www.ncbi.nlm.nih.gov/books/NBK560911/ doi:10.1016/B978-0-7020-7959-7.00042-7 PubMed PMID: 32809746.

36. World Health Organization. Diphtheria: risk communication and community engagement guidance [Internet]. 2026 [cited 2026 Apr 26]. Available from: https://www.emro.who.int/cpi/publications/diphtheria-risk-communication-and-community-engagement-guidance.html

